# Views and experiences of Every Mind Matters, Public Health England’s adult mental health literacy campaign: a qualitative interview study

**DOI:** 10.1101/2022.11.10.22282154

**Authors:** Ruth Stuart, Prisha Shah, Rachel Rowan Olive, Kylee Trevillion, Claire Henderson

**Affiliations:** Mental Health Policy Research Unit, Health Service & Population Research Department, King’s College London, Institute of Psychiatry, Psychology and Neuroscience, London SE5 8AF, UK; Mental Health Policy Research Unit, Lived Experience Working Group, Division of Psychiatry, Faculty of Brain Sciences, University College London, London WC1E 6BT, UK

**Keywords:** public health campaign, mental health literacy, common mental health problems, anxiety, stress, low mood, sleep problems, severe mental illness

## Abstract

**Background:** Every Mind Matters is a publicly funded health campaign, launched in England in 2019, to equip adults to look after their mental health, and that of others, by offering online information about common problems: anxiety, low mood, sleep, stress. This study is one component of an independent evaluation of Every Mind Matters conducted by the NIHR Mental Health Policy Research Unit. Its aim is to explore individuals’ experiences and views of the Every Mind Matters campaign and website.

**Methods:** Four researchers, including three with lived experience of using mental health services, conducted 20, one-off, semi-structured, online interviews with a range of adult participants, including a sample of EMM users and a purposively recruited sub-sample known to have severe or long-term mental health conditions. The team took a codebook approach to the analysis of the transcripts and identified four main themes.

**Findings:** There was an expectation from the name Every Mind Matters that its advice would address everyone. Almost all participants had experience of mental distress and looked to Every Mind Matters for help with a current problem for themselves.

All participants were complimentary about the Every Mind Matters website and found it to be user-friendly (theme 1) and personalised (theme 2) especially the interactive feature ‘Your Mind Plan quiz’ which responds with suggested actions to improve wellbeing and follows up with reminder emails. A few participants found this life changing.

Some participants wanted Every Mind Matters to better acknowledge the contexts in which they live (theme 3) such as the limitations of health conditions and health services, and difficulties of crowded housing, social policy, and climate change. Many participants would like Every Mind Matters to do more (theme 4), offer more interactivity, more choice, more information about available treatments, and more stratified advice to cover more severe mental health conditions so that everyone is included.

**Conclusion:** The population that experiences common mental health difficulties is not separate from the population that has severe mental health problems. Every Mind Matters could continue and build on its success by addressing a wider range of needs.

## Introduction

### Background

Prior to the COVID-19 pandemic, the global prevalence of common mental health problems over the past 12 months was found to be 1 in 5 amongst adults ages 16-65 in 63 countries (1). In England, a nationally representative general population survey (2) estimated that in 2014 1 in 6 adults (16+) had a current common mental health problem, whether treated or untreated. In the first six months of the pandemic, internationally (32 countries), the prevalence of common mental health problems increased to 1 in 3 (3) and in the United Kingdom a survey conducted in April 2020 (during the first lockdown) (4) found 1 in 4 (27%) adults (16+) had clinically significant levels of mental distress. In seeking to address the extent of mental health need among the general population, the English public health agency – Public Health England (PHE)^1^ - implemented a government-funded mental health literacy campaign.

The concept of mental health literacy was introduced by Professor Anthony Jorm in Australia in 1997 (5) as “knowledge and beliefs about mental disorders which aid their recognition, management or prevention.” In 2000 Professor Jorm (6) went on to write “…the prevalence of mental disorders is so high that the mental health workforce cannot help everyone affected […] If there are to be greater gains in prevention, early intervention, self-help and support of others in the community, then we need a ‘mental health literate’ society in which basic knowledge and skills are more widely distributed.”

Every Mind Matters (EMM) is a web resource with accompanying social marketing campaign to improve mental health literacy amongst adults (18+) in England to ‘be better informed and equipped to look after their own mental health and help others’ (7) and hence to significantly impact on population mental health by increasing mental wellbeing and reducing vulnerability to mental health conditions. EMM was created by PHE with expert advice from Professor Stan Kutcher who has developed mental health literacy resources for schools in Canada (8) and many other places.

Kutcher’s four-point definition of mental health literacy is ‘1. Understanding how to optimize and maintain good mental health; 2. Understanding mental disorders and their treatments; 3. Decreasing stigma; 4. Enhancing help-seeking efficacy.’

EMM was piloted in the Midlands in October 2018. Analyses of pre- and post-pilot surveys and interviews prompted a change of emphasis away from offering information about ‘mental disorders’ to recommending actions to improve wellbeing. EMM was launched nationally in England in October 2019 with an initial target to reach at least one million adults in England over three years.

The social marketing campaign for EMM is promoted through social media, radio and television, and offers partner organisations (e.g., National Health Service (NHS), local authorities, health charities, universities, and employers) a range of targeted materials (e.g., wallet-sized, conversation-starter information cards) to distribute to the public (9). The campaign and materials direct people to the EMM website (https://www.nhs.uk/every-mind-matters/) (10) which has psychoeducational content about four common mental health problems (anxiety, low mood, sleep and stress), self-help recommendations, guidance for helping others with mental health needs, possible causes of poor mental health, and contacts for urgent support. The most publicised feature of EMM is ‘Your Mind

Plan’, a quiz which generates a personalised plan of actions recommended to ‘protect and improve mental health’ (7). PHE research at the development of EMM showed that members of the public currently experiencing low levels of mental health difficulties were most interested in and likely to use EMM resources. It was fortunate that EMM was set up in advance of the outbreak of COVID-19 and could respond by adding a Coronavirus section (e.g., ‘mental wellbeing while staying at home’ and ‘money worries and job uncertainty’).

The National Institute for Health and Care Research (NIHR) Mental Health Policy Research Unit (MHPRU) conducted an independent mixed-methods evaluation of EMM. Components include:

1. Measurement: Development of three new measures of mental health literacy (recognition, action, vigilance) (11) suitable for the content of EMM and use in general population samples.
2. Change in outcomes over time and their relationship to EMM awareness or use: Quantitative analysis of data from repeated bespoke surveys of nationally representative samples before and since the launch of EMM using the above measures and others based on the Kutcher definition of mental health literacy (12).
3. Economic evaluation: Analysis of the Health Survey for England 2019^2^ dataset to assess whether use/awareness of EMM moderates the relationships between wellbeing and help-seeking, and wellbeing and the costs of primary care use; and modelling using the repeated bespoke surveys to examine the cost-effectiveness of the campaign (improvements in health literacy and well-being).
4. Experiences of the EMM campaign and web resource: Qualitative interviews to explore individuals’ views of the campaign, use of the web resources, and any barriers they encountered.

Here we report on 4. the qualitative study. The aim of this paper is to highlight thematically the experiences and views of our participants and to discuss the implications for EMM and the evaluation of public health campaigns generally.

## Methods

### Research Team

The individual roles and contributions of the research team (CH, KT, RS, PS, RRO and TK) are detailed under Authors’ Contributions and Acknowledgements.

### Study Design

Twenty individual semi-structured qualitative interviews about EMM were conducted. Interviews were recorded and transcribed verbatim and analysed using a Codebook thematic analysis, with a mixed inductive and deductive approach. We follow COREQ reporting guidelines (13).

### Participant Selection and Recruitment

#### Study Sample

Two samples were recruited to the study. For Sample 1, eligible participants had to have visited the EMM website for their own mental health or for someone close to them. For Sample 2, participants had to be aware of the EMM campaign and have experienced severe mental health problems not covered by EMM (e.g., bipolar disorder, an eating disorder, major depressive disorder, OCD, ‘personality disorder’, or schizophrenia) for a period of three years or more and/or engaged with secondary mental health services. All participants had to be living in England; be aged 18 years or older; be fluent enough in English to be able to understand the study documentation and the interview; and be able to provide informed consent. Sample 2 was included following discussion with the MHPRU’s Lived Experience Working Group. The Group suggested the inclusion of this additional sample could importantly capture the views and experiences of those whose mental health needs extend beyond the “low-level” the EMM campaign expected to serve and more fully explore any unintended impacts, positive or negative, of this widely available public resource. Diversity was sought by selecting participants by age, gender, ethnic background, and location, and for a range of self-reported mental health problems.

#### Recruitment methods

Participants were recruited primarily via an expression of interest form that PHE added to the EMM website in April 2021. Sample 2 were recruited via a previous MHPRU study of mental health service users. Participants from the MHPRU study who had consented to contact for future research studies were emailed by a MHPRU colleague (MB) and invited to contact our study email address.

RS emailed people who completed the EMM expression of interest form and attached an information sheet, consent form, and demographic questions. There were separate information sheets for Samples 1 and 2, but the demographic questions were the same for everyone. Questions included standard demographic categories and whether respondents belonged to groups identified by PHE to be at particular risk for mental health difficulties. The form also asked: ‘Do you have a mental health condition or mental health conditions?’ and ‘Do you have experience of using services for mental health prior to visiting the Every Mind Matters website?’ Finally, the form asked, ‘Which part/s, if any, of the Every Mind Matters website have you looked at?*’* The research team offered £20 as a thank you for participation.

### Setting and Topic Guide

Interviews were conducted remotely because of the COVID-19 pandemic. Participants could choose to be interviewed via Microsoft Teams, Zoom or a free telephone number linked to Zoom. Facilities on those platforms were used to record consent and then, separately, the interview. Interviews were conducted at a time when participants could not be overheard or interrupted. No one else accompanied the participants at the interview.

Sample 1 participants were interviewed by RS or PS. Sample 2 participants were interviewed by interviewers with lived experience PS, RRO and TK. Researchers introduced themselves and explained that they were independent from PHE. When not interviewing, RS was present (with camera and microphone off) to record all interviews. Individual interviewers decided whether to disclose their own lived experience depending on how the interview was going and whether they thought it would put the participant at ease.

PS conducted the first Sample 1 and Sample 2 interviews as pilots. No revisions were needed. Many of the questions and prompts were the same in the topic guides for each sample [see Additional files 1 and 2], but the topic guide for Sample 2 did not presume engagement with the EMM website and acknowledged that the participant had a mental health condition.

Interviews took place between 22 April 2021-12 October 2021. They ranged in length from 28 - 78 minutes. They were transcribed by a professional transcription service, then checked by the research team for accuracy. Transcripts were not sent to participants for comment or correction.

### Analysis

KT (Senior Lecturer in qualitative research) advised the study team. We took a codebook approach(15) which suited a small team. This approach is distinct from those which use a coding frame to consider concepts such as inter-coder reliability or data saturation and retains qualitative methodologies’ focus on researcher subjectivity.

To start, the team (KT, PS, RRO, RS, TK) each closely read and annotated the same two transcripts, one from each sample, and met to compare annotations and develop a set of descriptive labels/codes. The initial coding frame in NVivo Pro12 (16) included these inductive codes and some deductive codes based on questions asked in the interviews. The initial coding frame was used by researchers to code the transcripts of interviews they had personally conducted, so the reading of the texts was enriched by their recollections of the interviews. During coding five additional codes/sub-nodes were added as needed and agreed by the team. Analysis of the coded data was subsequently split between three researchers (RRO, PS, RS) based on their agreement about how different sections of the data related to potential themes. They each summarised sections of the data and drafted the related themes, and meetings were held to develop the themes. Writing of the Results was shared. The team includes researchers with lived experience of mental health difficulties and valuing their insights has been key to our analysis.

## Results

Participant ID numbers are in brackets. Those in Sample 1 begin ‘1.’ e.g., (1.1), and Sample 2 ‘2.’ e.g., (2.243).

### Sample

RS sent study information to 504 people. We received 49 replies, including 10 from men. One interview with a man had to be abandoned because he had no recollection of EMM. Seven people were excluded due to living outside of England and others withdrew for a range of reasons (e.g., ‘*overwhelmed’* by our 10 pages of study information and forms, technical difficulties, or bereavement). Two respondents were seeking help for their mental health. Eleven people responded to the email invitations for Sample 2, including three men.

Fourteen Sample 1 interviews and six Sample 2 interviews were arranged. (N.B. Two of the Sample 2 participants came via the EMM website. They reported their mental health diagnoses on the study demographic form and agreed that the Sample 2 interview questions would be more suitable for them.)

Table 1 sets out the demographic information about our two samples. A good spread of ages was achieved and five people with ethnicities that are in a minority in England were interviewed. Two English regions were unrepresented: Northwest England and Yorkshire & Humber.

**Table 1.** Sample Demographics.

|  | Sample 1 | Sample 2 | Combined sample N = 20 |
| --- | --- | --- | --- |
| female | 12 | 4 | 16 |
| male | 2 | 2 | 4 |
| 25-34 years old | 1 | 2 | 3 |
| 35-44 | 2 | 3 | 5 |
| 45-54 | 4 | 1 | 5 |
| 55-64 | 4 | - | 4 |
| 65-74 | 1 | - | 1 |
| 75-84 | 1 | - | 1 |
| 85-94 | 1 | - | 1 |
| Black Caribbean | 2 | 1 | 3 |
| Indian | 1 | 1 | 2 |
| White British | 11 | 3 | 14 |
| prefer not to say ethnicity | - | 1 | 1 |
| Northeast England | 2 | 1 | 3 |
| West Midlands | 1 | - | 1 |
| East Midlands | 4 | 4 | 8 |
| Southwest England | 1 | - | 1 |
| London | 2 | 1 | 3 |
| Southeast England | 3 | - | 3 |
| prefer not to say region | 1 | - | 1 |
| City > 100,000 | 4 | 5 | 9 |
| Small city or town | 8 | - | 8 |
| Village | 1 | 1 | 2 |
| Countryside | 1 | - | 1 |

Table 2 shows the samples’ characteristics based on PHE’s list of risk factors for the common mental health problems covered by the EMM website.

**Table 2.** Groups of Interest.

|  | Sample 1 | Sample 2 | Combined Sample N = 19<sup>a</sup> |
| --- | --- | --- | --- |
| Do you consider yourself as a carer? | 2 | 2 | 4 |
| Do you have a long-term health condition? | 4 | 6 | 10 |
| Are you a parent of a child/young person under 25 years old? | 4 | 3 | 7 |
| Are you LGBTQI? | 3 | 2 | 5 |
| Do you consider yourself as having a disability? | 1 | 5 | 6 |
| Have you been in contact with the Criminal Justice System in the last 12 months? | 2 | - | 2 |
| Are you currently struggling with debt? | 3 | 2 | 5 |
| Are you in unstable employment or unemployed? | 4 | 4 | 8 |
| Are you having housing problems? | 2 | 2 | 4 |
| Are you experiencing poverty, deprivation, or debt? | 1 | 2 | 3 |
| Have you experienced trauma? | 8 | 4 | 12 |
| Do you feel socially isolated? | 7 | 4 | 11 |
| Have you been divorced in the last 12 months<sup>b</sup> | 1 | 1 | 2 |
| Have you been bereaved in the last 12 months<sup>b</sup> | - | 1 | 1 |
| Prefer not to say. | - | - | - |

<sup>a</sup>data missing from one Sample 1 participant.
<sup>b</sup>The divorce and bereavement categories were added by PHE after this study had started. Four of the Sample 1 participants and one Sample 2 participant received an earlier version of the Demographic Questions and were not asked these questions.

In answer to the question about mental health, eight Sample 1 and all six Sample 2 participants noted one or more of the conditions/diagnoses listed in Table 3. One Sample 1 participant did not answer this question, and five reported no mental health condition.

**Table 3.**
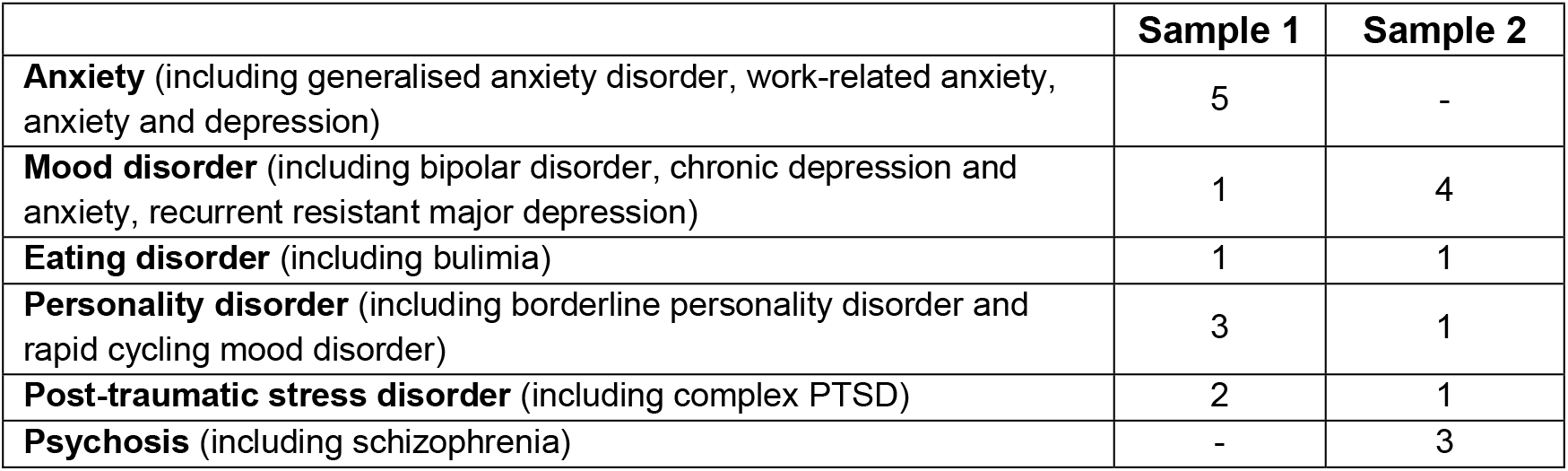
Mental health diagnoses as reported by participants on study demographic form.

|  | Sample 1 | Sample 2 |
| --- | --- | --- |
| **Anxiety** (including generalised anxiety disorder, work-related anxiety, anxiety and depression) | 5 | - |
| **Mood disorder** (including bipolar disorder, chronic depression and anxiety, recurrent resistant major depression) | 1 | 4 |
| **Eating disorder** (including bulimia) | 1 | 1 |
| **Personality disorder** (including borderline personality disorder and rapid cycling mood disorder) | 3 | 1 |
| **Post-traumatic stress disorder** (including complex PTSD) | 2 | 1 |
| **Psychosis** (including schizophrenia) | - | 3 |

Based on this overlap between samples 1 and 2, the same coding frame was applied to interviews from both samples. The overlap highlights the range of difficulties experienced by those who independently found their way to EMM via a range of referral routes.

### Descriptive Findings

#### How did people first hear about EMM

All participants were asked how they first heard about EMM. A few had seen advertisements on Facebook, Twitter or television. Two participants said they heard of EMM on the radio, but it was unclear whether these were advertisements or part of a discussion.

One participant heard about EMM on an online forum set up by a charity that supports people with bipolar disorder. Some discovered EMM via other NHS webpages, or recommendations from their General Practitioner, hospital or mental health service. Others heard about EMM through a staff wellbeing meeting, university health and wellbeing course notes, a Neighbourhood Watch newsletter, or advice accompanying a national COVID survey. It is unclear how many of these leads used dissemination materials provided by the EMM campaign.

One Sample 2 participant first heard about EMM from the invitation to this study. Others from both samples were searching online for help when they came across EMM.

#### Reasons why people sought out the EMM website

None of the participants were new to the subject of mental health. Almost all who looked at the EMM website were in search of immediate help for themselves, or for an ongoing need. Some were dealing with accumulated difficulties which originated from past trauma such as domestic violence or the unexpected death of a close family member. Others had sleep problems or recurrent physical health problems that exacerbated anxiety. Several participants spoke about stress at work or problems connected to the pandemic as front-line workers, or being on furlough, redundancy, or self-isolation. Many said that EMM was the only website they had visited for their mental health, but others had visited websites of various mental health charities, some for specific difficulties such as eating disorders and others providing more general support.

#### Methods used by people to access EMM

Most participants viewed EMM on their smart phone. Others used a tablet, laptop, or computer instead of, or as well as, a smart phone. Most had no technical difficulties. One thought EMM was ‘*a bit slow’* on their phone, and others attributed slowness to the age of their own devices.

#### Number of visits to EMM website

The number of visits to the EMM website varied. A few visited just once or twice. Most looked three to seven times over periods of three days to two years. Two engaged with the resource regularly amounting to more than 30 or 50 times:

> *“…probably six months […] When my mood has been quite low, I’ve gone on it more. But at least once or twice a week just to recap and sort of find some focus*.*”* (1.237)

Most participants said they would recommend EMM to others.

### Interpretive Findings

The analysis team (PS, RRO, RS) identified four main themes in the data: User-friendliness, EMM as a personal experience, Acknowledgement of contexts, and Users wanting more. Each theme is presented with sub-themes.

#### Theme 1 - User-friendliness

The creators of EMM have got a lot right. Participants liked the presentation of EMM, and they described a number of features that made the EMM website easy to use. These are organised under sub-themes: Visual aspects, and Language and tone.

##### Visual

Feedback on the visual aspects was universally positive, even from participants who were unwell at the time they were looking at the website:

> *“it was very easy to use. The page was easy on the eyes and I’m very happy to use the page again*.*”* (2.243)

There was a high regard for the use of appealing colours and fonts, a layout that was easy to navigate, and good use of headings and lists.

> *“the bold colours sort of like draw your attention to different things [*…*] the presentation’s great to be quite honest*.*”* (1.237)
>
> *“it is a very well-designed site, and it has lots of very useful menus and it helps you keep track of where you are and so it’s quite easy to go back and navigate*.*”* (2.2)
>
> “*Stick with your present designer!”* (1.200)

Information on the website is presented in small chunks interspersed with photographs and illustrations. Some people mentioned the embedded videos:

*“I think the main thing that helped me was that you have a choice to look at a video instead of reading lots of information*.*”* (1.710)

Another participant pointed out the benefit of videos for people who understand but cannot read English. However, videos did not suit everyone:

*“…one thing I really like is a good combination of visual and verbal statement… It’s your signs of anxiety screen. And it’s got a lovely list, a heading and a list of bullet points. Now I can take that in incredibly quickly. Whereas having to watch a video, I feel quite infantilised and irritated*.*”* (1.200)

The video clips have voice overs and music, but no participant mentioned anything related to the sounds on EMM.

##### Language and tone

Nearly all participants were positive about the quantity and quality of the text in EMM. They described the language as clear, succinct, easy to understand, and friendly without jargon:

> *“…like a snapshot of the illness. It gave you just the right amount of information*.*”*(2.10)
>
> *“…information is […] in quite straightforward terms, there isn’t really sort of any stigma attached to it, they don’t use unhelpful like or scary terminology like they don’t, it’s not overly medicalised*…*”* (2.59)

However, a few were concerned about the tone for young people:

> *“…the box about self-care for young people and how ‘2020 was pretty tough and 2021 has not exactly been all sunshine and rainbows either’ […] it feels like it’s sort of diminishing the issue because of the informal language used*.*”* (2.2)
>
> *“…there seems to be an assumption that it’s completely abnormal to be worried or anxious at all […] You have many positive, helpful suggestions, but there’s also this tendency to create a feeling - particularly around young people - that somehow if they’re not happy then they either have to blame themselves or somebody else*.*”* (1.200)

#### Theme 2 - Every Mind Matters as a personal experience

In recalling various aspects of their use of EMM, many participants spontaneously described how personal the experience was, particularly Your Mind Plan. Additional sub-themes comprise: EMM Emails, Swap Ideas, Structure, EMM as companion, and Change as a result of using EMM.

##### Your Mind Plan

Your Mind Plan is a quiz which asks five multi-choice questions about an individual’s mood, sleep, anxiety, stress, and what they might have been worried about over the past two weeks. It responds with a personalised action plan of six suggestions such as ‘Move more everyday – try a 10-minutes workout’. Participants reported that it was effective at engaging them. Only one participant did not start the quiz and almost all completed it.

> *“…initially it was just like take a quiz, which is quite a good way to go into something. You know, it draws you in quite nicely really”* (1.701)

For a participant who reported no personal mental health needs the quiz was reassuring:

> *“I think it was just more curiosity more than anything…So that’s probably why I’ve not gone back to Every Mind Matters, because it sort of summed it all up after the quiz and there wasn’t really anywhere for me to go next that I needed help with*.*”* (1.22)

A couple of others spoke about doing the quiz a number of times to monitor their health:

> *“And then when I feel like my mental health is not in a good place, then I’ll do the quiz to see, you know, where I am at…But I don’t do the mind quiz all the while, […] I do it when I’m feeling different*.*”* (1.237)

Not everyone felt the framing of the Your Mind Plan questions was right:

> *“…it really only looks at things within a two-week timeline but again that’s not very useful for people like me who, where the issues and feelings that we’re having are chronic and ongoing*.*”* (2.2)

##### EMM emails

EMM gives respondents the option to receive a copy of the Your Mind Plan action plan by email. Not everyone was aware of this option:

> *“You know, if I see something on the internet that I want to go back to, I’ll pin it. But the problem was, it went back to sort of home, just to advertising the plan. So I never got to keep my original plan. I screenshotted it. But […] I wasn’t aware at that point that I could have it emailed to me*.*”* (1.710)

One participant had problems because action plan email went into their Junk inbox.

> *“I was very put off by the fact that I didn’t get them […] And because of my mental health conditions, you now, it’s a vicious circle […] it sends you in a downward spiral. Every little thing like that, because everything makes me anxious*…*”* (1.701)

For people who opt to receive the action plan by email, EMM also generates follow-up emails as reminders or with extra advice (e.g., three weeks later, five months later, and at new campaign time points). This was highly appreciated by many participants.

> *“…having the email prompts that were asking “How are you getting on with the plan?”, “Have your goals changed?” eventually prompted me to actually follow through and I ended up following the plan […] I love them. I wasn’t expecting the follow-up emails, but it felt really good because a lot of the resources that I’ve seen, they just put the information out there and you can download a PDF or something, and that’s the end of it. But actually, having regular emails that are checking in on you and making sure that you haven’t forgotten the plan was really, really useful. And I felt like the website was a bit more personal and interactive than other resources that I’ve used*.*”* (1.62)

##### Swap ideas

EMM also gives respondents the option to edit the plan. Each suggestion has a ‘Not for you? Swap idea’ button which, when clicked, immediately substitutes the original suggestion with an alternative (e.g., ‘Try a 10-minute workout’ could be replaced by ‘A little activity every day’). The alternatives can be swapped too (e.g., ‘A little activity every day’ for ‘Get the couch to 5k app’ or ‘Get moving outside’). Again, participants found it helpful.

> *“…although it gives you a suggestion, you can change it if you want to. And that’s quite empowering, you know, you’re not being dictated to [*…*] you do have a choice*…*”* (1.1)

Some participants did not try the Swap function either because they were happy with the suggestions they were offered or because they were ambivalent about trying it.

> *“I did notice it. I wasn’t sure. I have mixed feelings on whether that was a good idea or not […] if you’re given, like, a prescription, as it were, and then you pick and choose out of those which ones helps you I don’t know how helpful that is going to be*.*”* (1.701)

A few participants from Sample 2 perceived the Swap function as unhelpful due to both the initial and subsequent suggestions offered not being perceived as helpful.

> *“… I think again when I used it, it swapped from try a mindfulness exercise to something like go for a walk or drink a cold glass of water which was equally not really the sort of help I was after*.*”* (2.2)

##### Structure

A couple of participants described how Your Mind Plan helped them implement change:

> *“…it gave me some practical advice that was a bit more long-term. It wasn’t just to keep me calm for right now. It was like ‘OK, this is what you can work on for the week’ […] Every Mind Matters […] gave me the structure that I needed to make […] stuff work […] it gave me some direction which was useful*.*”* (1.62)

They recognised the value in being able to tailor the Plan to their needs and be active agents in the personalisation:

> *“You can adjust it according to you, you can customise it according to your own needs […] it doesn’t tell you […] I can personalise it to my own, like if I like a certain activity, I can focus on more of that, and if I don’t…So that’s really helpful*.*”* (1.2)

##### EMM as companion

A number of participants personified EMM and seemed to feel supported beyond what might be expected purely from the informational content:

> *“Every Mind Matters, it’s almost like a buddy […] it’s not like a website that I’d never go to again. I feel they’ve done a profile for you and it’s almost personal now*.*”* (1.126)
>
> *“I think the website feels as though it’s a partnership with you […] it’s trying its best to help you*.*”* (1.1)
>
> *“…when I read through your website it brought a warm feeling to us because I thought, ah, here’s people that care, that are like willing to listen*.*”* (1.597)

Several participants reported feelings of reassurance that EMM was available throughout the day:

> *“I feel like it’s […] at hand 24/7 […] Sometimes when I get my isolation feeling where I don’t want to go out and I don’t want to do this, it’s there at hand if I need it*.*”* (1.237)

Having the resource readily available helped some participants feel less alone and more hopeful:

> *“Maybe it’s changed the thing that you’re not alone. […] mental health can isolate you and it has isolated me. And I have lost quite a lot of friends because of my mental health. […] So it’s helped me deal with that …*.*”* (1.237)
>
> *“…from years and years of getting tired with so called help, this website has renewed my hope that I will understand myself and … start the process of seeking that help again. But in the right direction*…*”* (1.710)

##### Change as a result of using EMM

For many participants EMM appeared to be a springboard for more confidence, action, or improved wellbeing.

> *“… since looking at the Every Mind Matters, I do feel a little bit better about going to university, and I started looking at joining the societies… it has made me a little bit more focused to combat my anxiety…”* (1.535)
>
> *“… before the Mind Plan, my headspace wasn’t sort of in the right place, you know, I wasn’t relaxing, I couldn’t focus. And since I’ve been on the website and done the Mind Plan, it’s really helped with my moods and helped me to stay focused and positive*.*”* (1.712)
>
> *“That’s what your page brings to people who are starting to feel depressed. You put a light at the end of the tunnel”*. (1.597)

Another participant reported that they did not follow through on the suggested actions but that it was validating to receive the ideas anyway:

> *“Well, it was interesting to look over and see that the sort of latest advice endorsed the sort of things that I have learnt and found myself over time. I was open to suggestions for when I feel anxious or negative. And it kind of cheered me up in that sense. I thought you’re doing quite a good job to be fair! So yes, it was useful*.*”* (1.200)

Similarly, some people described the advice offered as not new to them, but revisiting it as a result of the Mind Plan was helpful:

> *“So, for me, that was good to go back to and revisit because sometimes it wasn’t always new, what it emailed me, but it gave me the opportunity to think Oh, yes, I’ve done that in the past and that worked, and that revisiting which can be so beneficial*.*”* (2.10)

A couple of participants were thrilled by the results they achieved within a month with EMM. Both were practising the same two Mind Plan suggestions: Relax your muscles and your mind and Reframe unhelpful thoughts.

> *“Relax your muscles and your mind […] it’s been really, really useful, really useful. And the other one I have found also profoundly revolutionary is Reframing unhelpful thoughts [*…*] I’m just sticking with […] these couple of ideas but keep using them until it becomes much more natural to me. […] I can feel the change happening inside my head where there will come a point, you know, like learning to drive, where you don’t have to think anymore. It’s gone into your subconscious a bit. And that little tool then, hopefully, will sit there forever…”* (1.1)
>
> *“… for me, it’s quite significant. It’s de-demonising stress and making it something that, it’s a disability really that you can actually do something about. […] it’s certainly made a big difference and I’m, you know, more than chuffed […] So things have changed, my thinking has changed. […] I’m actually welcoming the challenge when stress arises […] Rather than panicking…”* (1.126)

A participant who had used EMM for several months explained how they made major steps. Shortly before the research interview they had spoken to their manager, were signed off work, and signed up for therapy.

> *“I’ve hid it for so many years and now I am honest that I have got a problem and I need to deal with that problem. Because it’s not going to go away. […EMM] points out, yeah, this is, this could be depression, this could be stress, this could be anxiety. So it gives you a pointer in the right direction and then, you know, sitting back in the early hours thinking, well, I’ve got that, I’ve got that and I’ve got that. Maybe I need to get some help with that. It’s helped me admit that I need some help*.*” (1*.*237)*

#### Theme 3 - Acknowledgement of contexts

Participants across both samples reported a desire for EMM to pay more attention to the contexts in which they experienced difficulties with their mental health. This data was divided into the following sub-themes: Personal contexts; Socio-political contexts; Health system contexts; and the context provided by previous campaigns.

##### Personal contexts

Personal contexts were the living situations of our participants, the individual circumstances which affected whether and how people were able to engage with suggestions the site made for self-help.

There was a degree of frustration that psychological and practical barriers to implementing such suggestions were largely unacknowledged.

> *“… firstly, it doesn’t know that I already do those things, that I’m already helping myself in every way that I know how, […] but equally then when it tells you to do something you did an hour ago, you think well then there’s no hope for me, if that’sthe answer and I’ve already done it and I still feel like this, then there’s no hope for me. […] it sort of put all the responsibility for my feelings and experiences on me, and it wasn’t about my condition, or it wasn’t about my circumstances […] Oh, you can help yourself in this way, and actually you can’t always. Sometimes you get beyond that and sometimes you do that, and it still doesn’t help, and sometimes you just can’t do it in the first place […] I don’t think it recognises […] that can make people feel worse*.*”* (2.59)

Sometimes psychological and practical barriers combined to reduce accessibility further. On a purely practical level, some participants said some suggestions that the Your Mind Plan gave were impossible for people to implement due to personal or material circumstances:

> *“I’ve got serious physical health illness, so when it talks about exercise, it’s a really hard one because I can’t go for a 10-minute walk, I can only walk with crutches, or I use a wheelchair*.*”* (2.1)
>
> *“I can’t possibly do a workout or meditation I just didn’t like that, because I have my young boy sleeping in the same room and the practicalities of listening to a video*.*”* (1.71)
>
> *“…for me, the breathing exercise almost kind of like, ok, well, breathe, so I’ve got nine people, so where do it do it? It doesn’t give me anything that says ‘Can you try and block out the noise?’ It’s more of a given that you’ve got a special place. In our house, that’s pretty much the bathroom*.*”* (2.3)

Some, but not all, participants who reported practical barriers were able to use the swap function in Your Mind Plan to find options which were more suitable to their circumstances.

On a psychological level, participants from both samples reported that the same difficulties which had led them to access the site, particularly obsessive and/or anxious thought patterns, could make completing the Mind Plan quiz or implementing self-help suggestions impossible, for example:

> *“…it really felt as though I was putting in quite a bit of energy to kind of respond to the questions. But then, actually, my anxiety was fueled on: what’s the result going to be […] am I going to be able to handle the result, or will I be able to do what it tells me to do, or will it be just something that I’ve done before that’s not worked?”* (2.3)
>
> *“…particularly when you’re very anxious if you found something you think it might be a solution, but then actually becomes too much work to deal with it. You just give up on it*.*”* (1.701)

##### Socio-political contexts

A few participants pointed to the absence of references to environmental or socio-political contributors to mental distress. One question on the Your Mind Plan quiz is ‘Have you been worrying about anything? Choose as many as apply: Coronavirus, Personal life and relationships; Money, work and housing; Life changes and difficult times; Health issues; Traumatic events; Smoking, drinking, drugs or gambling; None of the above.’

> *“Quite frankly I’ve learned to cope with the effects of this [depression and anxiety], but what really drives me to desperation is (a) climate, the future that confronts us and the behaviour of this government. You know, I’ve been reduced to tears by some of the things they’ve done. And that should be included in your list of what upsets people, you know, and leads to distress*.*”* (1.200)
>
> *“It would have been nice to have actually just said, you know that resilience is great, but there are also some really bad environments where actually you need more than resilience to actually survive*.*”* (2.3)

A participant also spoke about overcrowded housing and the unequal impact of COVID on different ethnic communities in their city. They concluded by saying that having time and space for self-care “*is almost a privilege*” (2.3) and felt that the EMM website did not recognise the challenges.

##### Health system contexts

Participants brought their existing knowledge of and feelings towards the NHS to their EMM experience.

> *“I see my GP about normal problems not about my mind*.*”* (1.216)

Experiences for many were inevitably informed by trying to seek mental health support during the COVID-19 pandemic.

> *“… we know that the NHS is under tremendous strain. We know mental health services are under tremendous strain*.*”* (1.701)
>
> *“…with the exception of your page [EMM Coronavirus section], there’s very little there for people to read […] who are struggling, you know, only last thing is, phone your doctor up, but then you’ve got to wait three days for a phone call*.*”* (1.597)
>
> *“… people in secondary care who have been almost ignored throughout the pandemic, they don’t have anything*” (2.3)

Participants who found out about EMM via NHS information leaflets described feeling positively predisposed towards EMM due to its NHS affiliation, which had associations with being trustworthy and evidence-based:

> *“You think, well, if it’s in this NHS leaflet then it’s something that should be worthwhile. Whereas ordinary adverts are only doing it to sell whatever it is they’re selling…”* (1.216)
>
> *“I was thinking, well, this is from the NHS, they’ve thought this through. In theory, they’ve had experts who have chosen these so I would feel safe in using the NHS ones*.*”* (1.701)

However, where people had experiences of exclusion, neglect, or difficulty accessing NHS services, this could be compounded by the site. While some felt that the site lived up to the promise of its name, “Every Mind Matters”, and the universal healthcare offer implied by its NHS provenance, others did not:

> *“Because it says Every Mind Matters, I hoped that it would cover the full spectrum of mental health illness. Because to me, that’s what the title’s saying […] I’m at the more serious end of the spectrum and it feels like we’re not valued. I’m not valued. We’re kind of forgotten*.*”* (2.1)

The signposting function of the EMM site relies on the healthcare system being able to provide the services to which people were signposted. This was not always the case.

> *“Urgent mental health support, find your local 24/7 mental health crisis line’. Well, here I think it’s called the Community Mental Health team. I rang them and then it rang out for 20 minutes. So, I gave up … I didn’t want Samaritans because I wasn’t suicidal. I was overwhelmed with anxiety*.*”* (1.701)

Others were frustrated at being signposted to things they already knew about or had already tried:

> *“…when you click through to the urgent help thing, it just diverts you to another NHS page […] there isn’t actually anything on that page […] even the site it signposts you to is still signposting, like there isn’t any kind of, I don’t know, anything at all interactive or that you could directly connect to a service that might help you if you needed to […] you feel like you’ve clicked enough links to be saying I need some help. I need some help, click, click, click. And then it just tells you like, oh, you could phone the Samaritans. You know, thanks, I kind of worked that one out…”* (2.59)

Participants who expressed these feelings of being overlooked, excluded, or frustrated did not necessarily report that EMM had no merit or no useful parts, but were clear that there was potential for EMM to contribute to harm in a context where those with complex mental health needs did not always have those needs met in the NHS more broadly.

##### Previous campaigns

A participant who, years ago, had been detrimentally impacted by a campaign for physical health, spoke about public health campaigns generally. They were concerned that a little knowledge could be a dangerous thing.

> *“I think campaigns like Mental Health Awareness Day and stuff and the Every Mind Matters site, […] it’s such a spectrum of experience that using, you know, having some depressive symptoms and not feeling stigmatised because of it is one thing, but actually eradicating stigma for treatment-resistant depression or like in its extreme forms […] I actually think they might make that problem worse. Because people think they understand it and have a good grasp on mental health and how to improve your own wellbeing and actually that can lead to them […] failing to recognise that that isn’t applicable to everyone*.*”* (2.59)

Not everyone felt that a limited remit was harmful. The participant who described feeling ‘not valued’ (Health systems context above), when asked who they thought EMM was for, said:

> *“I think the Every Mind Matters is almost like a beginner’s guide to mental health illness. And there’s nothing wrong with that”* (2.1)

#### Theme 4 – Users wanting more

All Sample 1 participants were asked if they had any suggestions to improve EMM and Sample 2 participants were asked how they might re-design EMM. Several had no suggestions, in some cases because they thought the resource was already “brilliant” (1.2) and “explains everything” (1.712). However, as much as participants liked website’s digestibility and appreciated not being overloaded with information, many wanted EMM to do more. The following sub-themes were developed: More interactivity, Gaps, Inclusive, and More information.

##### More interactivity

Some suggestions were about allowing greater engagement with EMM such as “a search function” (2.2) and “information sheets you can print off” (2.10). Two participants wanted to be able to log the things they were doing with EMM:

> *“… one thing I would have liked would be to be able to track my progress. So, for example, with the five things that they recommended, if there was some way to keep a log of what I’d done and on what day…”* (1.62)
>
> *“… a before and after and a bit more of a timed approach”* (1.126)

A participant also suggested a little survey *“what do you find that is working?”* and more EMM emails asking, *“how are you getting on?”* or *“did you know about this feature?”* (1.126)

Another participant thought that EMM should encourage people who had swapped suggestions offered by Your Mind Plan *“to go back and look again at the things you turned down”* (1.701).

Other participants made suggestions either because they found gaps in the resource or because they wanted it to be more inclusive.

##### Gaps

During the interviews some participants mentioned things they noticed were missing from EMM: environmental factors and actual urgent support (Socio-political and Health system contexts above), and specific life events or issues which were not covered.

> *“I did find there was a lot of life changes there, which I’ve got, which aren’t listed* … *divorce is one of them*.*”* (1.701)

One Sample 1 participant intended to seek EMM advice for an elderly relative about dementia, which is not covered, and another thought there should be signposting for long COVID.

Participants who were frustrated with the Urgent support section offered suggestions to reduce this.

> *“…there should be something there and then for you to speak to somebody … you know it’s not a crisis … say if I woke up tomorrow and I was at university, but I was feeling anxious about going, if you could just contact someone, go on a web forum and explain how you’re feeling and someone could say, well I think that you should deal with it this way, that would help … Just a bit more support for people …. I think a lot more people would be less anxious and depressed if there was more help like that out there*.*”* (1.535)

EMM was accessible to this participant because it is free. They had found links to other help online which they could not use because it required payment.

Another participant recommended that the ‘Urgent support’ be formatted differently, rather than a section like others on the website.

> *“I don’t think it should look like that’s what the site is really offering because it doesn’t…”* (2.59)

This participant also recommended that Your Mind Plan quiz have a threshold at which respondents are advised to contact their GP.

##### Inclusive

It was clear that some participants in Sample 2 were disappointed by the EMM website:

> *“It just wasn’t what I’d hoped for*.*”* (2.59)
>
> *“I just think it could have been braver*.*”* (2.10)
>
> *“… if you’re somebody that actually goes through real mental illness, it’s not, there’s not enough depth there to actually deal with it*.*”* (2.3)

They wanted their chronic experiences and less common or more serious conditions to be acknowledged and covered by EMM too: eating disorders, self-harm (including for adults), PTSD, bipolar, schizophrenia, neurodivergence and autism.

> *“… it’s a very good site for people who are like experiencing their first struggles but not so much people with chronic conditions*.*”* (2.2)
>
> *“… the website just seemed as though it’s very much targeted to the majority of people and the minority, it’s almost like we don’t really exist. There’s no specific area or section for us on there …”* (2.3)

A Sample 2 participant made suggestions for more plurality: low-level and higher-level interventions to cover people in primary and secondary care. Similarly, alternatives to suit people in different circumstances. For example, the simple, abstract breathing exercise currently offered by EMM made one participant in the countryside feel “*secure*” and “*grounded*” (1.710), whereas a city-dwelling participant preferred something more immersive that would block out background noise and worries.

Other suggestions were to include *“Testimonies from people with mental health problems who’ve gone through it*.*”* (2.3) and to *“Help people see difficulty as a starting point*.*”* (1.200) and acknowledge that:

> *“…people relapse. It’s not a continuum, it’s a very zigzaggy line, mental health illness* …*”* (2.10)
>
> *“… people who are disabled by their mental health conditions […] you can still do stuff and just because someone’s at work, doesn’t mean they don’t have a disability …”* (2.59)

In the absence of coverage of more severe conditions, this participant recommended that EMM state its purpose and include a disclaimer:

> *“… if you’re already under the care of your local mental health team, consider contacting them rather than relying on this site …”* (2.59)

##### More information

Sample 2 participants had many constructive ideas for how EMM might develop.

One (2.2) suggested a “*glossary […] just for acronyms*”, “*area-specific information about local services*”, “*a section on medications*” and “*how to order online prescriptions*”, *“introductions to the various types of therapies offered on the NHS […] what they can do and where can they be accessed […] and whether or not they might be useful for you or whether you’d be eligible for them”*, all to:

> *“… facilitate a much more patient-led treatment system so that it’s possible to be more informed about the kind of treatment that you’re being offered and that you’re more empowered to make those decisions in an informed way*.*”* (2.2)

Another thought EMM could be more comprehensive and create a “*directory*”:

> *“…for example, psychosis […] it would give you a basic description what psychosis looks like or what it is. And then it would actually signpost you to relevant websites that could actually give you support. But they would all be sub-sectioned with history of psychosis, current treatments, wellbeing activities, maintenance, things like that […] targeted so that the website [EMM] would be a place […] where you could go to actually know where to go*.*”* (2.3)

## Discussion

The purpose of the study was to report people’s experiences and views of the EMM campaign and online resources. Participants discovered EMM through a range of routes. They were very complimentary about the design of the website: how it looked, ease of use, and its content. Most participants completed the Your Mind Plan quiz and welcomed the action plan it generated and follow-up emails. Many felt the suggested actions to be very personal and useful and structured in a way that was actionable. Several participants thought EMM should acknowledge the contexts to people’s mental health: the difficulties of health conditions and some people’s circumstances; the social and environmental problems many face; and the limitations of health services. Many participants would like EMM to do more: offer more personal interactivity, cover more conditions in more detail, and aim to be inclusive of everyone.

All participants appreciated the presentation and accessibility of EMM. The participants who experienced most difficulty using EMM were anxious and recognised that their anxiety disrupted their interaction with EMM. The repeated use of Your Mind Plan quiz as a tool for monitoring wellbeing was a user innovation. The enthusiasm for EMM’s 24/7 accessibility contrasts with the data we collected about participant’s previous experiences of seeking help for mental health, which was mixed and included many frustrations and unmet needs.

The study shows that EMM’s anticipated core audience of adults with common mental health problems is not separate from the population who have experienced severe difficulties and distress. More than half of the 14 Sample 1 participants recruited via the EMM website reported trauma, including attempted suicide, and more than half had a mental health diagnosis. With this level of lived experience in Sample 1, as well as in Sample 2, many of our participants already had a relatively high level of mental health literacy and their needs therefore differed from Kutcher’s audience in Canadian schools (8). Most of our participants were not seeking to ‘optimize and maintain good mental health’ but rather turned to EMM for help to manage problems they already knew about and struggled with for years. The study includes one clear example of EMM effectively aiding a participant to name their problems and seek professional help, and others who found the self-help suggestions recommended EMM’s Mind Plan to be life changing.

However, this mismatch between the anticipated core audience and the users we interviewed means that while there is potential for campaigns like EMM to support mental health very well, they are not risk-free. Public health campaigns aimed at populations with common mental health difficulties or at early intervention cannot in reality be targeted so that they are only seen by those groups. Public health campaigns are seen and used by the whole public, which includes people with more severe and long-term difficulties. Consequently, some participants felt ignored and disappointed by EMM content, particularly where its focus on early intervention and basic mental health literacy compounded experiences of exclusion and marginalization which existed elsewhere in people’s lives or mental health treatment.

### Strengths and Limitations

One of the strengths of the study is the depth of insight participants gave, which included positive, neutral, and negative experiences. Proactively contacting Sample 2 was another strength. Sample 2 participants who felt the EMM resource did not address their needs, nevertheless contributed many thoughtful suggestions about how EMM could develop and broaden the support it offers.

The recruitment method meant that the study only included people who self-selected to take part, but we sought to address some of these limitations by recruiting a purposive sample to ensure diversity among participants. The diversity of participants’ social and ethnic backgrounds is a strength of the study. It may be that including age categories up to 85-94 and >95 on the study demographic form attracted older participants, with decades of lived experience, to take part in a mental health research study where they are often not represented.

Despite some strengths in diversity, and despite the research team prolonging the recruitment period to increase participation among men, the study has an under-representation of men (4 out of 20). Only around 25%-30% of details submitted by respondents via the EMM website appeared to be from men, compared to around 70-75% from women.

Another limitation is that interviews were conducted on weekdays between 10:00-18:00. Some people may not have responded to the invitation for an interview because it clashed with their own working hours. Weekends and extended hours could be offered in future studies.

### Implications

Many participants, both those whose needs were or were not met, would like EMM to do more. Suggestions ranged from adding a search function, to an interactive online service where the same doctor or health worker would be available on a regular basis to offer free personal 1:1 advice. This points to the importance of evaluating EMM in the context of the broader UK healthcare landscape in which some people’s mental health support needs are often not being met (17) (18). It is one thing to establish a remit focused on common mental health difficulties and early intervention when there are meaningful resources to signpost, and another to try to establish that remit when those with long term or severe difficulties find their way to EMM and may have existing experiences of exclusion and unmet needs compounded.

Sample 2 participants who felt least served by EMM seemed most ambitious for its further development to support their inclusion. The integration of more stratified advice to include long-term manifestations of the conditions EMM already covers as well as more severe mental conditions could extend public literacy and understanding and possibly reduce stigma (19).

EMM could consider adding content for over-80s many of whom will have family or friends affected by various mental illnesses and dementia and could benefit from EMM’s companionable support.

In practice the audience for public health campaigns is any member of the public. Future research evaluation of public health campaigns and tools could consider seeking the views and experiences of those who may see the campaigns but who are not its primary core audience; as with EMM, this may reveal unintended impacts, positive or negative.

## Conclusion

‘Every Mind Matters’ is a promising and well-received title which drew in many of our participants. Participants in this study experienced the EMM website as user-friendly and personalised, but they wanted EMM to better acknowledge the contexts in which they live and would like EMM to do more. This suggests EMM should be continued and build on its success so it can become more inclusive.

## Data Availability

Although the interview transcripts have been pseudonymised, participants' individual circumstances may be recognizable. In order to protect participants' confidentiality, the interview transcripts will not be publicly available. The interview recordings have been destroyed.

## Lived Experience Commentary

The Every Mind Matters (EMM) website appears to be a helpful, interactive resource aiding people to increase their mental health literacy and offering personalised self-help tools. The free resources across a variety of categories appear to empower people with knowledge and tools to prevent their mental health deteriorating, and in some cases seek help.

Although the name ‘Every Mind Matters’ suggests the website is for every person, the site does not cater for people with diagnosed mental health conditions. It assumes people with a diagnosis have access to support through mental health services, which we know isn’t always the case. By shying away from talking about different mental health conditions and levels of risk, it is not for ‘every mind’ and promotes exclusion and ‘othering’ of those with diagnoses. To improve, EMM could add a disclaimer on the front page about who the website is and isn’t helpful for, or provide resources and information for different mental health conditions to ensure every mind who visits the website can access the support and resources they need. Furthermore, we would recommend inclusion of more material on Relationships and Postnatal depression.

The website automatically excludes people with limited computer literacy or those who are not in a position to afford a computer or a tablet, as well as those whose mobile phone cannot run such an application. It also doesn’t consider those whose English is not their first language. We would recommend having a translation feature on the website for prominently used foreign languages, and the same would apply for EMM leaflets available in GP surgeries or community centres. The developers have not catered for potential users with disabilities such as visual impairments or hearing difficulties who would struggle with the reading material or videos.

We have been impressed by the campaign’s user-friendly content, especially its smooth flow, simple language and font and just the right amount of information that doesn’t feel overwhelming, rather making the navigation more enjoyable. The practicality of ‘Your Mind Plan’ resulting from the Mind quiz is also commendable. By using the Mind plan, we were impressed by its being tailored towards the exact help needs expressed, which makes the whole experience personalised but also practical, giving hope for a positive change.

Elizabeth Mitchell and TK

## Abbreviations

APMS: Adult Psychiatric Morbidity Survey
DHSC: Department of Health and Social Care
EMM: Every Mind Matters
GP: General Practitioner
IAPT: Improving Access to Psychological Therapies
LEWG: NIHR MHPRU Lived Experienced Working Group
MHPRU: NIHR Mental Health Policy Research Unit
NHS: National Health Service
NIHR: National Institute for Health and Care Research
OCD: Obsessive compulsive disorder
OHID: Office for Health Inequalities and Disparities
PHE: Public Health England
PTSD: Post-traumatic stress disorder

## Declarations

### Ethical approval and consent to participate

The study was approved on 13 January 2021 by the Psychiatry, Nursing and Midwifery Research Ethics Subcommittee (PNM RESC) at King’s College London. Ethical Clearance Reference Number: HR-20/21-20911.

### Consent for publication

All participants received a copy of the approved consent form by email in advance and consent was recorded immediately before each interview. The interviewer read aloud the 11 statements and the participants consented, or not, to each in turn including ‘9. I agree to the use of anonymous quotations from my interview being used in research presentations and reports’. These five-minute recordings have been retained as proof of consent.

### Availability of data

Although the interview transcripts have been pseudonymised, participants’ individual circumstances may be recognizable. In order to protect participants’ confidentiality, the transcripts will not be publicly available. Interview recordings have been destroyed.

### Authors’ contributions

CH wrote the protocol and topic guides, and supervised participant selection and the whole study. KT co-wrote the topic guides and supervised coding and qualitative analysis. RS wrote the study documents and ethics application, recruited and selected participants, interviewed, coded transcripts, contributed to the analysis and interpretation of findings; co-wrote the Results and drafted the other sections of the paper. PS and RRO contributed ideas to the development of the project, including study documents; interviewed participants; contributed to the analysis and interpretation of findings; and co-wrote the Results. All authors contributed to and approved the final manuscript.

## Acknowledgments

We thank PHE colleagues for their collaboration; Mary Birken for circulating the Sample 2 invitation; TK for commenting on the topic guides, information sheets, demographic questions, and consent form, interviewing participants, and coding transcripts; and Sonia Johnson and Alan Simpson for their support as Directors of the NIHR MHPRU.

## Additional files

Additional-file-1-Every-Mind-Matters-Sample-1-interview-topic-guide.pdf

Additional-file-2-Every-Mind-Matters-Sample-2-interview-topic-guide.pdf

## Footnotes

Footnote 1 The Office for Health Improvement and Disparities (OHID), which replaced PHE in October 2021, is now responsible for EMM.

Footnote 2. The NHS Digital Health Survey for England monitors national health trends annually and estimates the proportion of people with health conditions and the prevalence of risk factors. https://www.gov.uk/government/statistics/health-survey-for-england-health-survey-for-england-2019 (14)

## References

1. Steel Z, Marnane C, Iranpour C, Chey T, Jackson JW, Patel V, et al. The global prevalence of common mental disorders: a systematic review and meta-analysis 1980-2013. Int J Epidemiol. 2014;43(2):476–93.

2. McManus S, Bebbington P, Jenkins R, Brugha T. Adult Psychiatric Morbidity Survey 2014 Executive Summary. Executive Summary. Leeds: NatCen Social Research in collaboration with the University of Leicester, commissioned by NHS Digital fbDoH; 2014 2016.

3. Nochaiwong S, Ruengorn C, Thavorn K, Hutton B, Awiphan R, Phosuya C, et al. Global prevalence of mental health issues among the general population during the coronavirus disease-2019 pandemic: a systematic review and meta-analysis. Sci Rep. 2021;11(1):10173.

4. Pierce M, Hope H, Ford T, Hatch S, Hotopf M, John A, et al. Mental health before and during the COVID-19 pandemic: a longitudinal probability sample survey of the UK population. The Lancet Psychiatry. 2020;7(10):883–92.

5. Jorm AF, Korten AE, Jacomb PA, Christensen H, Rodgers B, Pollitt P. Mental health literacy: a survey of the public’s ability to recognise mental disorders and their beliefs about the effectiveness of treatment. MedJAust. 1997;166(4):5.

6. Jorm AF. Mental health literacy. Public knowledge and beliefs about mental disorders. Br J Psychiatry. 2000;177:396–401.

7. Farrell S. Every Mind Matters. In: Care DoHaS, editor.: Public Health England; 2020.

8. Kutcher S, Bagnell A, Wei Y. Mental health literacy in secondary schools: a Canadian approach. Child Adolesc Psychiatr Clin N Am. 2015;24(2):233–44.

9. PHE. Campaign Resources-Better Health-Every Mind Matters: Public Health England; [cited 2022. Available from: https://campaignresources.phe.gov.uk/resources/campaigns/111-better-health---every-mind-matters/resources#.

10. England PH. Every Mind Matters England: Department of Health and Social Care; [Available from: https://www.nhs.uk/every-mind-matters/.

11. Chua K-C, Hahn JS, Farrell S, Jolly A, Khangura R, Henderson C. Mental Health Literacy: a focus on daily life context for population health measurement. Social Science & Medicine - Mental Health. 2022;2(100118).

12. Hahn JS, Chua K-C, Henderson C. The Every Mind Matters campaign in England: changes in mental health literacy over 30 months and associations between campaign awareness and outcome. medRxiv. 2022.

13. Tong A, Sainsbury P, Craig J. Consolidated criteria for reporting qualitative research (COREQ): a 32-item checklist for interviews and focus groups. International Journal for Quality in Health Care. 2007;19(6):349–57.

14. Mindell J, Robinson C. Health Survey for England 2019. In: Ucl, NatCen, editors.: NHS Digital; 2019.

15. Braun V, Clarke V. Can I use TA? Shoud I use TA? Should I not use TA? Comparing reflexive thematic analysis and other pattern-based qualitative analytic approaches. Couns Psychother Res. 2021;21:37–47.

16. QSR. NVivo 12 Pro. 2018 p. QSR International Pty Ltd.

17. Davies SC. Public Mental Health Priorities: Investing in Evidence. In: Health Do, editor. 2013.

18. Plewes J. Analysis: the rise in mental health demand: NHS Confederation; 2022 [07/09/2022]. Available from: https://www.nhsconfed.org/articles/analysis-rise-mental-health-demand.

19. Morgan AJ, Reavley NJ, Ross A, Too LS, Jorm AF. Interventions to reduce stigma towards people with severe mental illness: Systematic review and meta-analysis. J Psychiatr Res. 2018;103:120–33.

