## Additional file 1 for "Views and experiences of Every Mind Matters, Public Health England’s adult mental health literacy campaign: a qualitative interview study"

### Every Mind Matters interview topic guide for Sample 1

#### Welcome & explanation

Thank you for giving up some of your time to talk to me today. In this interview, I'll ask you about your views of the Every Mind Matters campaign, your experiences of using the website, any particular strengths and weakness of the site, what you did after using the site, and what you may do in future having used the site. The interview usually takes about 40 minutes but may be a bit shorter or longer depending on how much you want to say on each question.

#### Guidelines

There are no right or wrong answers. This is about your experiences and views.

Please remember that you can take your time to answer the questions, you do not have to answer any of the questions if you do not wish to, and you can take a break or terminate the interview at any time. You can withdraw completely from the study without needing to give a reason. If at any point you start to feel upset or distressed in any way, then just let me know and we can take a break or talk about something you find easier. You could also discuss any concerns at the end of the interview, when the recording has stopped.

#### Obtaining consent

[*Press record*]

Have you read through the information sheet about the study?
Do you have any questions before we begin? [*Interviewer answers any questions*]

Can I now ask you to please refer to the consent form. I need to record your consent. This will be a short recording separate to the main interview. Can I please ask you to confirm your consent to question 1, [continue reading questions 2-11 from qualEMM-consent-form-V4-25/02/21].

#### Introduction ending

Thank you. Do you have any other questions related to the interview?

[*Stop recording*]

I have stopped recording now and will start a new recording for the interview. Before I do, I just want to confirm that the interview recording will be transcribed and all identifying information such as names, places or services you have used, will be replaced by pseudonyms such as 'brother', 'city in NW England', or 'local psychotherapy service'. Following accuracy checks of the transcriptions, the recording of the interview will be deleted.

[*Press record*]

#### Interview

1. How did you first hear about Every Mind Matters (EMM)?

    a. Did you see/hear it advertised?
    b. Where? E.g. TV, GP surgery, gym?
    c. What did you think of the advertisement?

2. What made you want to use the EMM website?

    a. Before using the website, what were you expecting?
    b. Do you feel the website met your expectations?
    c. In what way?
    d. If not, why/how?

3. Please tell me about your use of EMM.

    a. What were your general impressions of the site and its content?
    b. How many times do you think you went onto the site?
    c. Can you say more about how you used the site?
    d. Which pages did you open?
    e. Did you try the Your Mind Plan quiz?
    f. (If yes) Did you use the Your Mind Plan action plan? How? E.g. Did you use some parts more than others? What made you engage more with some parts or not engage with other parts?
    g. Are you still using Your Mind Plan? How do you use it?
    h. Do you have any suggestions for how the site or action plan could be improved?

4. How did you find using EMM?

    a. Did you have any problems finding your way around the site?
    b. Did you use it as the phone app or on the website for PCs or tablets?
    c. Were there any technical problems?
    d. Were there any problems that meant you stopped using it?
    e. Did the content feel relevant to you?
    f. How did you find the presentation of the information?

5. How did using EMM make you feel?

6. Did using EMM change the way you think about anything?

7. Did you try anything new to change your mental health as a result of using EMM?

    a. How did you find it? Was it effective? Did EMM recommend anything you couldn't do, or didn't want to do? (Explore reasons behind this.) How did that make you feel/act?
    b. Did you use the 'swap idea' feature in your plan? How did you find this?

8. Do you plan to continue doing something for your wellbeing as a result of using EMM, or not?

9. If you were using EMM for a particular problem, please can you tell me a bit about this?

    a. When did it start?
    b. Had you ever had *x* before?
    c. (If yes) what if anything did you do about it previously?

    [This time] (if has had a previous episode) What if anything had you already done about x before you used EMM?

    a. Had you talked about it to anyone, such as friends or family?

b. (If yes) Can you say more about that? How helpful was that? What did they suggest? did you use this? Why/why not?/ What was doing this like? Did you find this useful? How did it help?
c. Had you seen a GP or another professional about x?
d. (If yes) Can you say more about that? How helpful was that? What did they suggest? did you use this? Why/ why not?/What was doing this like? Did you find it useful? How did it help?
e. Had you looked for other websites or apps?
f. (If yes) did you find anything online to try?
g. (If yes) Can you say more about that? What was doing this like? Did you find it useful? How did it help?

10. Did you do other things to help with x after using EMM or at the same time?

    a. (If yes) Please can you tell me about this?

11. Did using EMM help on top of the other things you tried, or not?

12. Have you had any other problems with mental health besides *x*?

    a. (If yes) what have you tried for help with this? (Probe for EMM.)

13. Did you look at any information specifically relating to the Coronavirus pandemic on the EMM website? Did you find it helpful / relevant to your experience of the pandemic? In what way(s)?

14. Who if anyone do you think EMM is helpful for? Why?

15. Have you recommended it to anyone?

    a. (If yes) Please tell me a bit about them without saying their name
    b. (If no) Would you recommend it to anyone you know or not? Why?

16. Are there any other points you'd like to raise about your experiences and views on EMM?

[*Stop recording*]
