## Additional file 2 for "Views and experiences of Every Mind Matters, Public Health England’s adult mental health literacy campaign: a qualitative interview study"

### Every Mind Matters interview topic guide for Sample 2

#### Welcome & explanation

Thank you for giving up some of your time to talk to me today. In this interview, I'll ask you about your views of the Every Mind Matters campaign and your experiences of using the website. The interview usually takes about 40 minutes but may be a bit shorter or longer depending on how much you used Every Mind Matters and want to say on each question.

#### Guidelines

There are no right or wrong answers. This is about your experiences and views.

Please remember that you can take your time to answer the questions, you do not have to answer any of the questions if you do not wish to, and you can take a break or terminate the interview at any time. You can withdraw completely from the study without needing to give a reason. If at any point you start to feel upset or distressed in any way, then just let me know and we can take a break or talk about something you find easier. You could also discuss any concerns at the end of the interview, when the recording has stopped

#### Obtaining consent

[*Press record*]

Have you read through the information sheet about the study?
Do you have any questions before we begin? [*Interviewer answers any questions*]

I now need to record your consent. This will be a short recording separate to the main interview. Can I please ask you to confirm your consent to Q1, [*continue reading questions 2-11 from qualEMM-consent-form-V4-25-02-21].*

#### Introduction Ending

Thank you. Do you have any other questions related to the interview?

[*Stop recording*]

I have stop recording now and will start a new recording for the interview. Before I do, I just want to confirm that the interview recording will be transcribed and all identifying information such as names, places or services you have used, will be replaced by pseudonyms such as 'brother', 'city in NW England', or 'local psychotherapy service'. Following accuracy checks of the transcriptions, the recording of the interview will be deleted.

[*Press Record*]

#### Interview

1. How did you first hear about Every Mind Matters (EMM)?
    a. Did you see/hear it advertised?
    b. Where? e.g. TV, GP surgery, gym?
    c. What did you think of the advertisement?
    d. Who do you think the advertisement was aimed at?
    e. How did the advertisement make you feel anything about yourself/your own mental health?

2. Have you used or are you still using the Every Mind Matters website?

[*For participants who did not use the site, skip to Question 4*]

3. What made you want to go to the Every Mind Matters website?
    a. Before using the website, what were you expecting?
    b. Did the website meet your expectations?
    c. If yes, in what way?
    d. If not, how?

4. If you didn't use the site, what put you off?
    a. Was it something about the way Every Mind Matters was advertised?
    b. Was it something in your own life that created a barrier (e.g. time, internet access, energy, your own mental or physical health)?

[*For participants who did not use the site, skip to Question 8*]

5. Please tell me about your use of Every Mind Matters.
    a. What were your general impressions of the site and its content?
    b. How many times do you think you went onto the site?
    c. Can you say more about how you used the site?
    d. Which pages did you open?
    e. Did you try the Your Mind Plan quiz?
    f. [*If yes*] Did you use the Your Mind Plan action plan? How?
    g. Did you have it emailed to yourself?
    h. Are you still using Your Mind Plan? How do you use it?

6. Technically, how did you find using the Every Mind Matters website?
    a. Did you have any problems finding your way around the site?
    b. Did you use it as the phone app or on the website for PCs or tablets?
    c. Were there any technical problems?
    d. Were there any problems that meant you stopped using it?
    e. Did the site suggest you download an app, or more than one app? Are you still using of these follow-up activities?

7. Was the Every Mind Matters website content relevant to you?
    a. What did you think of the way the advice is presented on the Every Mind Matters website?
    b. How did the site make you feel about yourself / your mental health?
    c. Are there any situations in which you would use the site again?
    d. Might the advice on Every Mind Matters have been more relevant to you at an earlier stage of your life?

8. Did using EMM or seeing the adverts change the way you think about anything?
    a. Did you try anything as a result of using EMM?
    b. How did you find it? Was it effective?
    c. Did it recommend anything you couldn't do, or didn't want to do? [*Explore reasons behind this.]*
    d. Did you use the 'swap idea' feature in your plan? How did you find this?

9. Do you plan to try continuing doing something for your wellbeing as a result of using EMM, or not?

10. How did the guidance on the Every Mind Matters site (or adverts) fit with other mental health treatment / advice you have received from mental health services

    a. Were any of the Every Mind Matters suggestions (or information) new to you?
    b. If you were using EMM for a particular problem, please can you tell me a bit about this?
        - When did it start?
        - Had you ever had x before?
        - [*If yes*] what if anything did you do about it?
        - Had you looked for other websites or apps?
        - [*If yes*] did you find anything online to try?
          [*If yes*] Can you say more about that? What was doing this like? Did you find it useful? How did it help?

11. Who if anyone do you think EMM is helpful for? Why?

12. Have you recommended it to anyone?

    a. [*If yes*] Please tell me a bit about them without saying their name
    b. [*If no*] Would you recommend it to anyone you know or not? Why?

13. If you were asked to re-design the Every Mind Matters website, would you:

    a. Add advice/address any mental health problem that isn't there now?
    b. Exclude any content currently on the site?
    c. Change the way the content is presented?
    d. Leave things more or less the same?
    e. If you would not want to redesign the campaign but would rather have a completely different mental health campaign (not necessarily limited to online content) what would that look like?

14. Are there any other points you'd like to raise about your experiences and views on Every Mind Matters?
